# Case Study: Using Facebook Data to Monitor Adherence to Stay-at-home Orders in Colorado and Utah

**DOI:** 10.1101/2020.06.04.20122093

**Authors:** Ryan Layer, Baily Fosdick, Michael Bradshaw, Dan Larremore, Paul Doherty

**Affiliations:** University of Colorado, 1111 Engineering Drive, Boulder, CO 80309

**Keywords:** COVID-19, population density, social distancing

## Abstract

In the absence of effective treatments, social distancing has been the only public health measure available to combat the COVID-19 pandemic. In the US, implementing this response has been left to state, county, and city officials, and many localities have issued some form of a stay-at-home order. Without existing tools and with limited resources, localities struggled to understand how their orders changed behavior. In response, several technology companies opened access to their users’ location data. As part of the COVID-19 Data Mobility Network, we obtained access to Facebook User data and developed four key metrics and visualizations to monitor various aspects of adherence to stay at home orders. These metrics were carefully incorporated into static and interactive visualizations for dissemination to local officials.

All code is open source and freely available at https://github.com/ryanlayer/COvid19

## 1. INTRODUCTION

On March 24th, 2020, at 1800 MNT, our team started collecting data from Facebook Data for Good [1] that covers Utah and Colorado. For these regions, Facebook provides aggregated and anonymized user density data for 2km × 2km tiles at three time points per day (0200 MNT, 1000 MNT, and 16000 MNT) for the eight hours following the time point (Figures 1 and 2A). We also began recording higher resolution data (0.6km × 0.6km tiles) for Boulder County, CO and the City and County of Denver, CO on March 30th and April 7th, respectively. To protect privacy, Facebook does not report density data for tiles with fewer than ten users. In addition to the current density data (crisis density), Facebook provides a baseline density for each tile and time point which is averaged over the same day and time periods during the 45 days before the start of data collection. Facebook Data for Good provides other types of mobility data, including movement between tiles, but the combination of Facebook’s privacy policy and the prevalence of large rural and remote areas in Utah and Colorado limited the utility of these data for our purposes.

**Figure 1.**
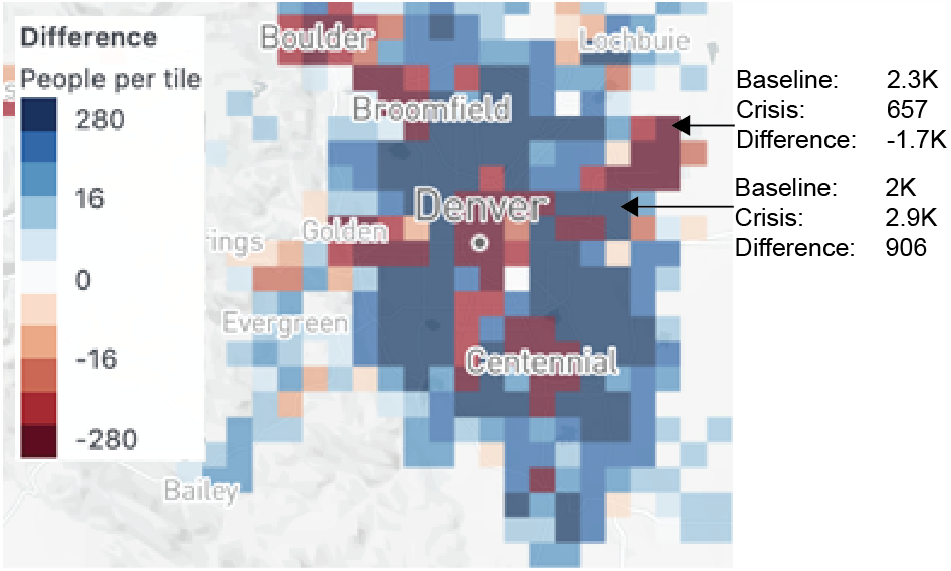
Facebook Data For Good map of Denver, CO on April 12, 2020, at 10L00 MNT. Cells are colored by difference between the current (crisis) and baseline user density data.

**Figure 2.**
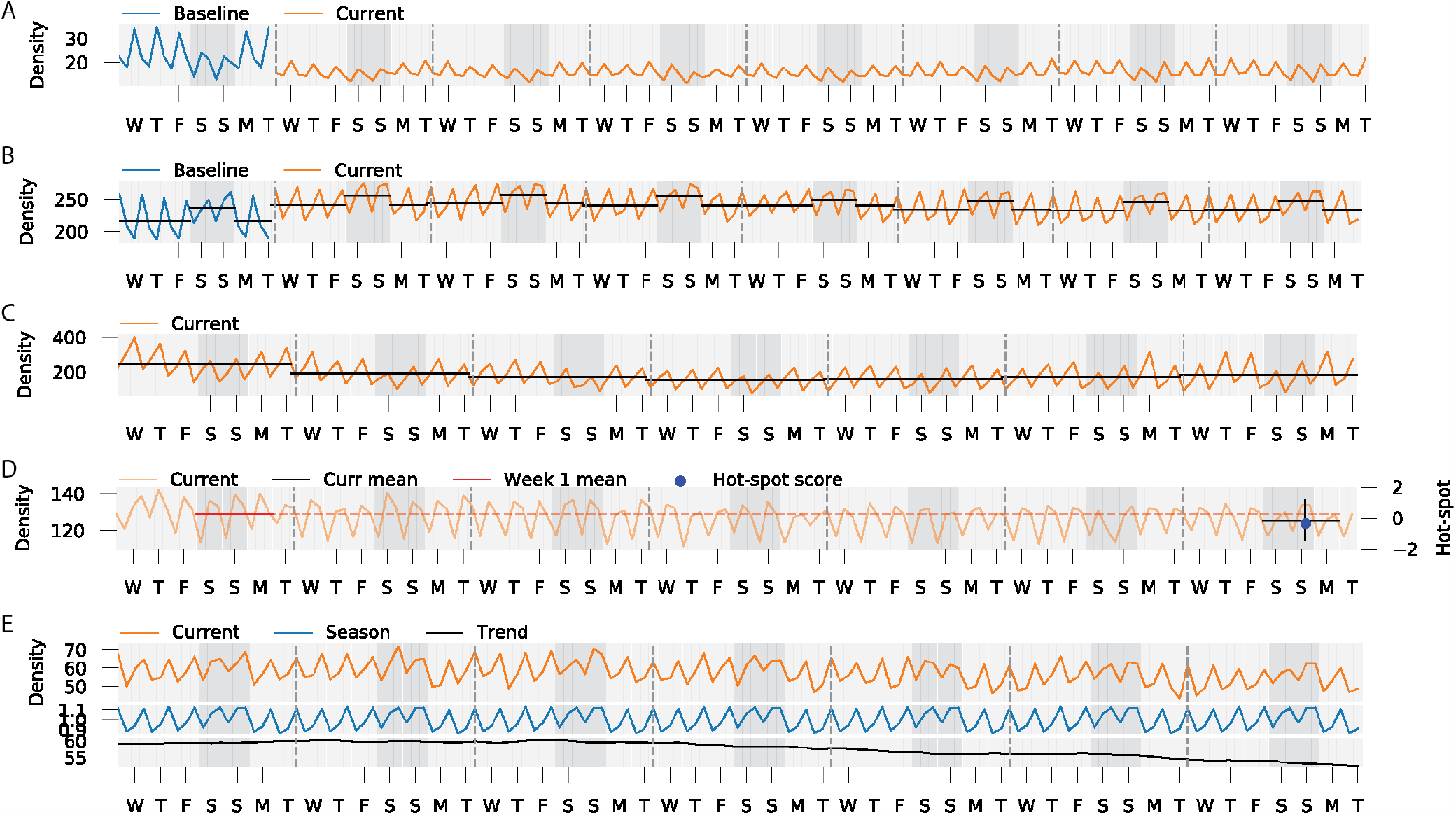
A) Baseline and current user density tracks for an individual cell in the Colorado map. Each day (shaded grey) includes time points at 0200, 1000, and 1800. Weekends are darkened. B) The weekend score compares the weekday mean (black bar in light grey shade) to the weekend mean (black bar in dark grey shade). C) The slip score compares adjacent week means (black bars). D) The hotspot score (blue dot) compares the current mean (horizontal black bar), the first week mean starting on the same day (red), and the variance in similarly dense cells (vertical black bar). E) Current densities (orange) are decomposed into the seasons (blue) and trends (black).

Governor Polis of Colorado mandated a statewide stay-at-home order starting Thursday, March 26, which shifted to a safer-at-home order on April 27. While Utah never enacted a statewide directive, Salt Lake County began a “Stay safe, Stay home” order on March 30, 2020 and relaxed down to a “Stay smart, Stay safe” order on April 17. As regulations continue to be relaxed, efficient monitoring is needed more than ever to assess the impacts of new policies on population movement.

## 2. METRICS

Cities and neighborhoods have unique dynamics and interpreting behavior from density fluctuations can be difficult. Before stay-at-home orders were issued, most tiles followed a general weekend to weekday pattern where weekday density was greatest in business districts and weekend density was greatest in residential areas. After social distancing policies started, these patterns were disrupted and comparing current behavior to baseline behavior often gave counterintuitive results. For example, adherence to stay-at-home orders in some residential areas appeared to be poor based on density fluctuations when in reality the density changes were a result of an overall increase in population by about 5%. We hypothesize that such increases were due to the return of college students and a drop in business travel, both of which are good social distancing practices. Given these nuances, we developed metrics with the base assumption that compliance starts out high and degrades over time. These metrics are targeted at monitoring transitions to and from stay-at-home orders.

### 2.1 Weekend Score

Under normal circumstances, most regions show a regular weekly pattern. Economic centers are denser on the weekdays, and residential areas are denser on the weekends. Under a stay-at-home order, the differences between weekend and weekday activity in regions containing non-essential businesses should be minor.

To measure weekday and weekend density differences, we developed the weekend score (ws) that compares the average weekday density dwd to the average weekend density dwe during a week:

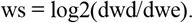

Areas with higher positive values have more weekday activity and areas with lower negative values have more weekend activity (see Figure 3A).

**Figure 3.**
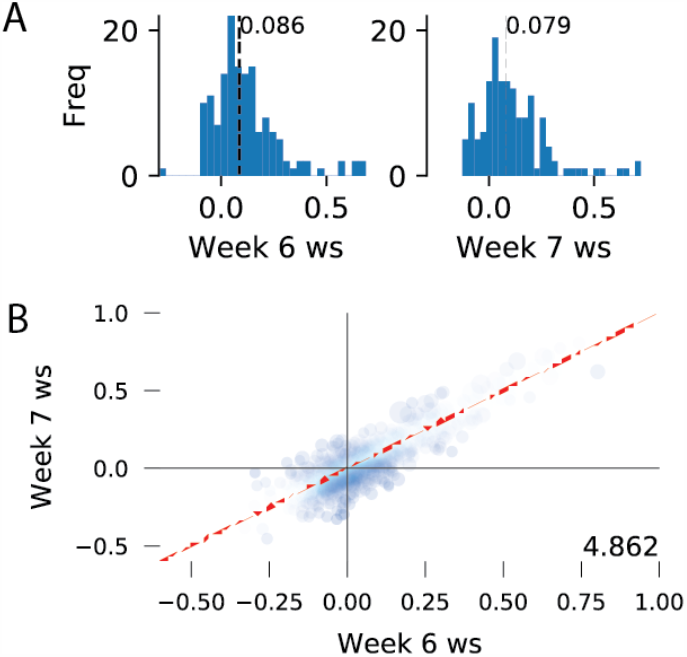

We track changes in behavior by comparing the distribution of scores between weeks or by the change in the behavior of individual tiles (see Figures 4 and 5). In Figure 5, each point is a tile and its size corresponds to the mean baseline density. Points further to the right or higher indicate more weekday activity. The closer a point is to the diagonal red line, the more similar the activity between the two weeks. We identify weeks based on the collection start date so “week 1” is effectively the first week of the stay-at-home order. This plot also shows a change from the baseline toward more consistent behavior between the weekdays and weekends.

**Figure 4.**
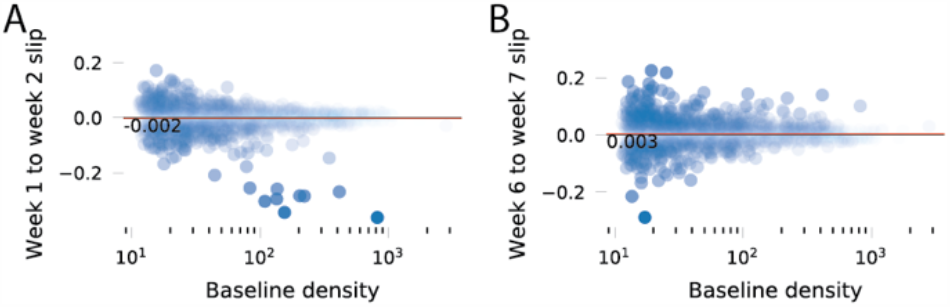

**Figure 5.**
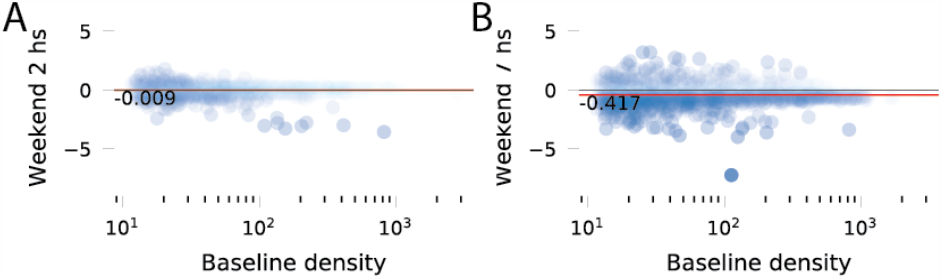

### 2.2 Slip Score

The behavior of a region after a stay-at-home order is issued depends on the region’s zoning and demographics, which complicates monitoring. For example, economic centers will be less dense, and residential centers will be denser since most people are not working or traveling. To track how well regions are adhering to their new patterns from week to week, we developed a slip score (ss). This metric assumes that adherence to the stay-at-home orders is best immediately following their issuance, then tracks changes by comparing the average weekly density di of consecutive weeks (see Figure 6):

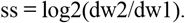

**Figure 6.**
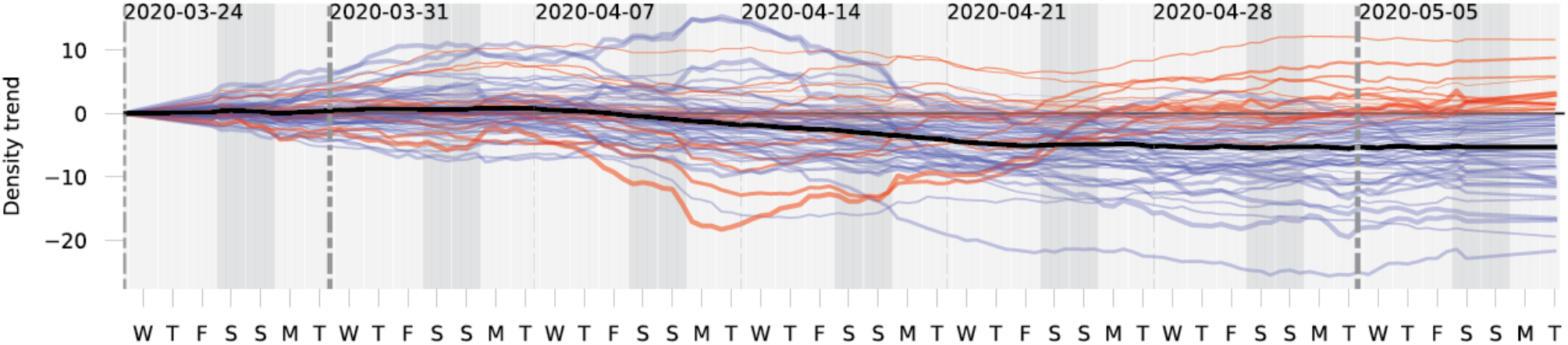

Areas with higher positive values were more active in the second week, and areas with lower negative values were less active.

Equal changes in density counts result in smaller slip scores in more populated tiles so we visualize slip score with respect to the region size. In Figure 7, each point is a tile, and points from denser regions are to the right are. Points above the zero line have slipped into a more active state, and points below the line have become less active.

### 2.3 Hot Spot Score

While the weekend score quantifies behavior pattern changes within weeks and the slip score captures gross density changes across weeks, we found local officials desired a finer resolution metric to assist in identifying sudden changes in population behavior. As a result, we developed a hot spot score (hs). This metric resembles a z-score as it is equal to the current 3-day density average d3_c_ minus the corresponding 3-day average from week 1 d3_w1,_ divided by an estimate of the standard deviation of the 3-day averages:

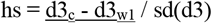

Here we are again assuming the first week of the stay-at-home order represents the state of highest compliance and present the current mobility patterns relative to that. Density patterns are averaged over 3-day windows to prevent spurious single-day fluctuations for receiving too much attention. In other words, the score for Monday, for example, is an average over densities observed on Saturday, Sunday, and Monday.

The purpose in scaling the density difference in the numerator by a standard deviation was to create a z-score, such that scores across tiles of different densities are comparable. However, the optimal method of estimating the standard deviation of a tile’s 3-day average is unclear. Using the given tile’s 3-day averages across the observed time period, e.g. all Saturday-Monday averages, results in an estimator that is based on fewer than six data points per tile and hence extremely variable. Furthermore, a tile that experienced large swings in density would have a large tile standard deviation and a low hot spot score, which is undesirable. Since denser tiles typically have larger variability, we estimated the standard deviation for each tile and 3-day window, fit a linear regression to these standard deviations as a function of average tile density, and used the fitted values from the model in the calculations of hot spot scores.

### 2.4 Trends

Density data can be represented as a time series with repeating short-term patterns (seasons) and the overall value increases or decreases (trends). Using a model, we can decompose a series into its seasons and trends [2], which provides a means to compare tiles with different weekday/weekend activities (seasons).

Even within a city, we can observe the differences in density dynamics between the business centers, which should be less dense during a stay-at-home order, and residential centers, which should be denser. To capture the overall trend of a city, we take the mean of each tile’s trend weighted by the tile’s baseline density.

## 3. DATA PRODCUTS

Governments are hierarchical, and the process of synthesizing complex data into recommendations is often performed by analysts and presented to policymakers. The availability and expertise of these analysts vary widely among local governments. To accommodate different data needs, we produce a range of products at different levels of granularity.

### 3.1 Weekly Situational Awareness Reports

Our most refined product is a one-page weekly situational report that includes a bulleted summary of the previous week’s activity and graphical views of our metrics. The sitrep is ideal for offices that do not have the capacity to interpret the metrics we provide. Even for localities with robust analytics capabilities, the weekly sitrep is a useful artifact that can be easily passed up to policymakers and shared across agencies.

### 3.2 Interactive Data Browser

### 3.3 ArcGIS Maps

Geographic information system (GIS) is a framework for storing and visualizing information spatially that allows analysis to interpret data by combining different information sources as layers on top of local maps. Many cities and counties have invested heavily in GIS and employ highly trained analysts that can combine their technical expertise with their local knowledge to interpret complex data sources quickly. To leverage these resources, we provide generated shapefiles that contain scores for each tile and publish these shapefiles as an ArcGIS feature.

## 4. CONCLUSIONS

Understanding the behavior of a city or county from population density data and integrating that knowledge into public health decisions is an evolving problem. We have focused on making close personal connections with local decision-makers to ensure that we are collecting the most insightful metrics and delivering the most useful data products. These connections will be vital as public health priorities shift, and data needs change.

Comments reflecting the utility of this work from our local government partners are given below:

CCD intends to utilize the data to evaluate the relationship between mobility and COVID-19 Case Rates by geographic area, e.g. does higher mobility lead to higher case rates and vice-versa. We may also use the data to inform our social distancing/safer-at-home order enforcement activities. [SOURCE]

In lieu of widespread testing and the data that comes from it, the Arapahoe County Office of Emergency Management is building an early warning system of areas that could become hotspots with the potential to overwhelm the medical and public health systems. Density and mobility are key indicators in this customized early warning system. We infuse the mobility data from these reports into a larger system that helps anticipate areas of potential viral spread, thus allowing for a more proactive response in regards to PPE supply distribution, and targeted community testing. [SOURCE].

I have found the sitreps useful for supplementing information I am getting from a few other sources to help me provide updates to our agency leadership and external partners about what we are seeing in terms of social mobility and changes over time. [SOURCE]

While our metrics and visualizations have been developed specifically for our local partners, we believe they will be either directly useful to other localities or can serve as starting points for more specific data needs.

## Data Availability

All code is open source and freely available at https://github.com/ryanlayer/COvid19

## 5. ACKNOWLEDGMENTS

We would like to thank the team from Facebook Data for Good, including Alex Pompe and Mercedes Erdey for helping us understand, access, and debug the density data. And our government partners: Nathan Fogg from Arapahoe County Sheriff’s Office, Office of Emergency Management; Mark Mullan, Megan Noel, Megan Hatten, Kristen Daly, Emily Paye, Audrey Schroer and Carol Helwig from Boulder County Public Health; Andrew Blunk and Paul Kresser from the City and County of Denver Technology Services; and Ilene Risk, Michelle Dallon, and Nicholas Rupp from Salt Lake County Public Health.

## About the authors

John Q. Miner is the Data Mining Guru at DM University, where he leads a life of decadence and excess. He currently manages strategic directions in the industry. Previously, he worked in ATU. He earned a Ph.D. from the Computer Science Department of Epicurian University, where his research was supported by a National Science fellowship. (www.lustforlife.com/~johnm)

**Author 2** is blah blah balh…

## REFERENCES

[1] Facebook Data For Good. https://dataforgood.fb.com/.

[2] Skipper S. and Perktold J. statsmodels: Econometric and statistical modeling with python. Proceedings of the 9th Python in Science Conference (Austin TX, June 2010).

